# Assessing capacity to social distance and neighborhood-level health disparities during the COVID-19 pandemic

**DOI:** 10.1101/2020.06.02.20120790

**Authors:** Daniel Carrión, Elena Colicino, Nicolo Foppa Pedretti, Kodi B. Arfer, Johnathan Rush, Nicholas DeFelice, Allan C. Just

**Affiliations:** Department of Environmental Medicine and Public Health, Icahn School of Medicine at Mount Sinai, New York, NY; Institute for Exposomic Research, Icahn School of Medicine at Mount Sinai, New York, NY

## Abstract

The COVID-19 pandemic has yielded disproportionate impacts on communities of color in New York City (NYC). Researchers have noted that social disadvantage may result in limited capacity to socially distance, and consequent disparities. Here, we investigate the role of neighborhood social disadvantage on the ability to socially distance, infections, and mortality. We combine Census Bureau and NYC open data with SARS-CoV-2 testing data using supervised dimensionality-reduction with Bayesian Weighted Quantile Sums regression. The result is a ZIP code-level index with relative weights for social factors facilitating infection risk. We find a positive association between neighborhood social disadvantage and infections, adjusting for the number of tests administered. Neighborhood infection risk is also associated with capacity to socially isolate, as measured by NYC subway data. Finally, infection risk is associated with COVID-19-related mortality. These analyses support that differences in capacity to socially isolate is a credible pathway between disadvantage and COVID-19 disparities.

## Introduction

The 2019 novel coronavirus (SARS-CoV-2) emerged in Wuhan, China, and has since become a worldwide pandemic. In the United States, given the nature of this novel infectious disease, anyone exposed to the pathogen was believed susceptible to infection, there were no proven pharmacologic treatments, and testing capacity was low. Pre-existing conditions are known risk factors of disease severity, and mortality increases sharply with age^1^. Consequently, the United States federal, state, and local governments have principally relied on non-pharmaceutical interventions such as social distancing and mask-wearing. New York State (NYS) on PAUSE is one such effort, whereby essential workers, i.e. healthcare workers, food purveyors, bank tellers, etc., were the only employees that should be reporting to work^2^. We examine the role of social factors, such as employment and commuting patterns, population density, food access, and personal finances and access to healthcare, in infection risk.

It has been widely noted in popular media and emerging scientific evidence that COVID-19 is taking a disproportionate toll on communities of color^3–6^. For example, in Chicago, Blacks comprise 70% of COVID-related deaths, but only 30% of the population^6^. In New York City (NYC), Hispanics/Latinx and Blacks are disproportionately impacted, representing 34% and 29% of the deaths, but 28% and 22% of the population, respectively^6^. While differences in disease severity are likely attributed to higher levels of preexisting conditions, i.e. health disparities^7^, this does not explain differences in disease incidence. A survey of laboratory-confirmed hospitalized cases across 14 states found, where race was reported, that 33.1% of hospitalized patients were non-Hispanic Black^8^. In NYC, as of May 13, 2020, the cumulative incidence of non-hospitalized positive cases were 798.2, 684.8, and 616.0 per 100,000 for Blacks/African Americans, Hispanic/Latinx, and Whites respectively^9^.

A body of literature on the social determinants of health suggest that there are numerous inequities that provide the scaffolding for increased COVID-19 infection rates in communities of color. Racism operates on both the interpersonal and structural levels, the latter explaining the societal mechanisms that reinforce inequality, including through housing, employment, earnings, benefits, health care, criminal justice, etc.^10^. Those structural forms of social disadvantage are responsible for many of the health disparities we observe in communities of color^11^.

Researchers have outlined the ways in which residential segregation and structural disadvantages lay the groundwork for racial disparities in infectious diseases^12^. More recently, others have noted that social distancing is more difficult for communities of color^6^. Taken together, this literature highlights the social mechanisms that facilitate viral spread in communities of color. The underlying structural disadvantages relevant to the current coronavirus pandemic might include that people of color (POC) are more represented amongst low-wage jobs^13^, many of which are now deemed essential^14^. When they get home from work, they are more likely to return to densely populated homes and neighborhoods^15^. Further, multigenerational homes are more common in communities of color^16^, making social distancing between least susceptible (healthy children) and most susceptible (elderly adults with chronic conditions) difficult. POC often live further from supermarkets and sources of nutritious foods, necessitating further travel for groceries^17^. These factors, among others, underscore the many ways that the capacity to social distance may be contextual, and based on structural factors.

In this study, we use socioeconomic data on neighborhood characteristics to understand differences in infection incidence between neighborhoods, as we quantify the relative contribution of these measures of social disadvantage and if a proxy of social isolation, NYC subway utilization, helps us to understand these differences. We create a ZIP code level infection risk index for NYC and show how this index explains racial/ethnic disparities in cases, thus reflecting structural forms of disadvantage. Finally, we examine the relationship between neighborhood infection risk and neighborhood-level COVID-19 mortality. Ultimately, we create a tool that identifies social factors that facilitate viral spread, and therefore, may be useful throughout the US to pinpoint potential areas for targeted public health intervention.

## Results

### Cross-sectional neighborhood infection risk index

We wanted to identify any association between a neighborhood social disadvantage composite index and cumulative COVID-19 infection incidence. There were 174,614 positive tests across 177 NYC ZIP Code Tabulation Areas (ZCTAs) as of May 7, 2020. Kendall’s tau correlations between social disadvantage variables ranged from −0.15 to 0.61. Kendall’s tau correlation tests were also conducted between each variable and the infection incidence (**Supplemental Table 2**). An assumption of the Bayesian Weighted Quantile Sums (BWQS) regression is that the direction of the effect for each variable is the same as the overall effect. Given the a priori hypothesis that increased disadvantage yields higher infections, we used the reciprocal of variables that are negatively associated with the infection incidence.

The BWQS regression analysis identified evidence of an association between our composite variable of ZCTA-level social disadvantage (on a ten unit scale) and the number of infections per 100,000 (**Figure 1**). We found that each unit increase in social disadvantage is associated with a 10% increase in infections per capita (Risk Ratio: 1.10; 95% Credible Interval: 1.08, 1.11). While all included variables contributed to this composite, they do not all contribute equally (**Figure 2**). We found that the average number of people in a household is the single largest contributor, followed by the proportion of the population who are essential workers and rely on personal vehicles or public transit to commute. Proportion of uninsured and the median income are also relatively informative compared to the other variables.

**Fig. 1.**
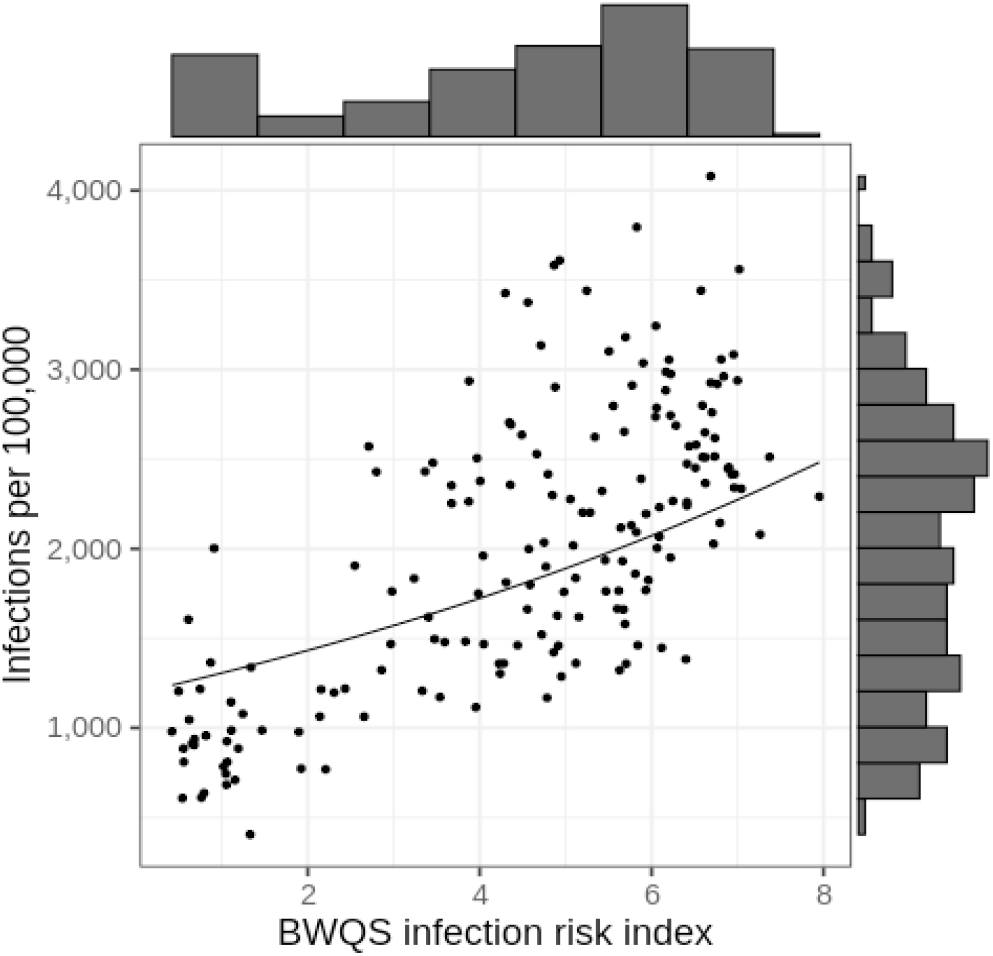
Scatterplot of BWQS infection risk index and cumulative infection incidence. Unit of analysis is ZCTA (n=177). Fitted line represents the BWQS regression line, holding testing ratio constant at the median, and marginal histograms represent the distribution of the variable on each axis.

**Fig. 2.**
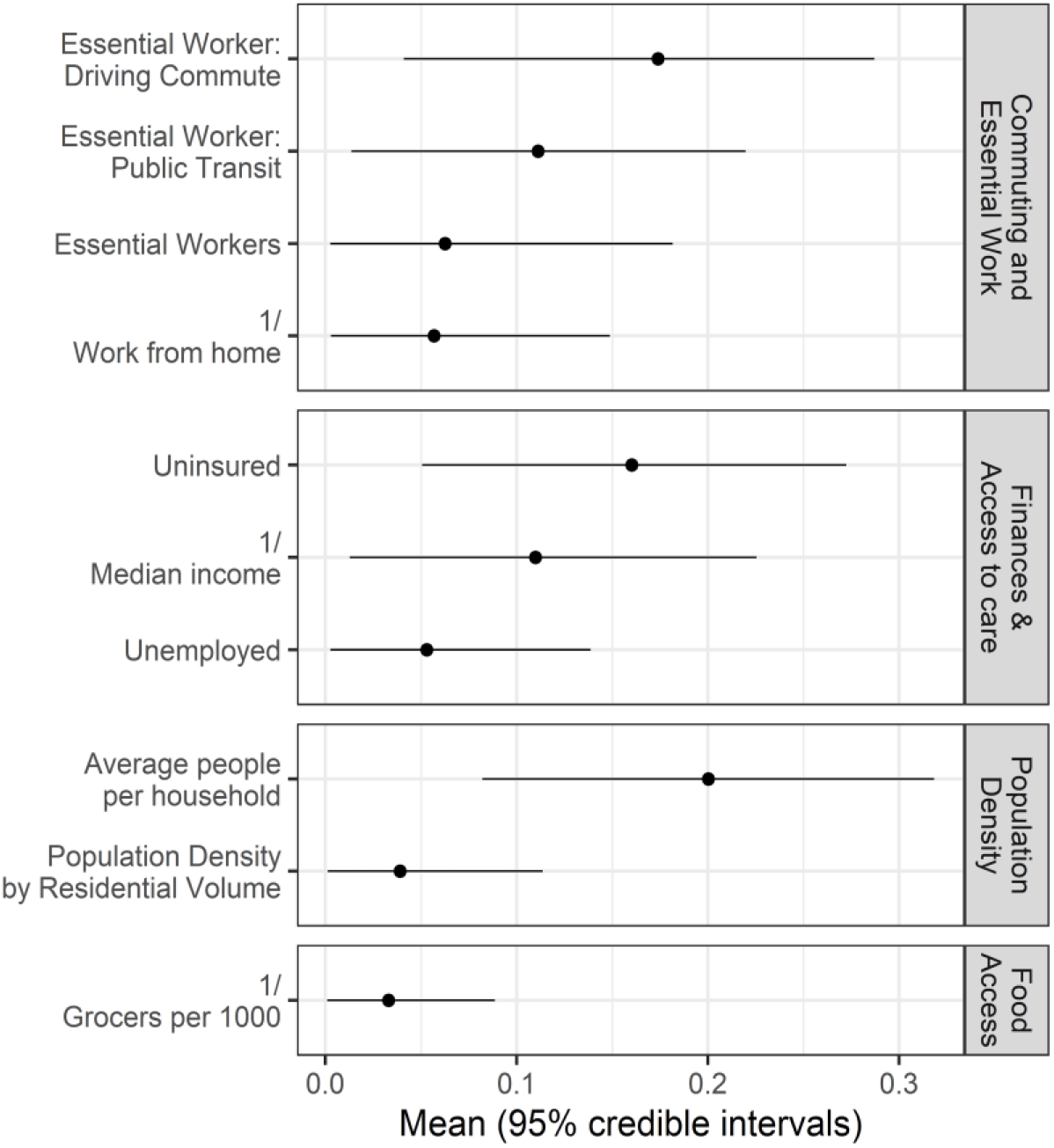
Estimated contribution of social variables to the BWQS infection risk index, with 95% credible intervals. The BWQS weights the explanatory variables by their relative contribution to the composite infection risk index, between 0 and 1. The mean weights sum to 1 and are organized into conceptual domains and ordered by mean weight.

The spatial distribution of the BWQS infection risk index (**Figure 3**) largely mirrors that of infections in NYC (**Supplemental Figure 1**). We examined the population demographics of neighborhoods according to their BWQS infection risk index (**Figure 4**). The data shows that Blacks have the highest population-weighted mean index and Whites have the lowest. Examining these distributions by quantile of the BWQS Index shows that White populations are overrepresented in ZCTAs in the lower quartile of the infection risk index (<25th percentile) and underrepresented in the upper quartile of infection risk (>75th percentile) ZCTAs (**Supplemental Figure 2**). While Whites comprise approximately 32% of NYC’s population, they only make up 11% of high infection risk ZCTAs. Conversely, Blacks and Hispanic/Latinx are 22% and 29% of NYC’s population and 31% and 42% of high risk areas respectively.

**Fig. 3.**
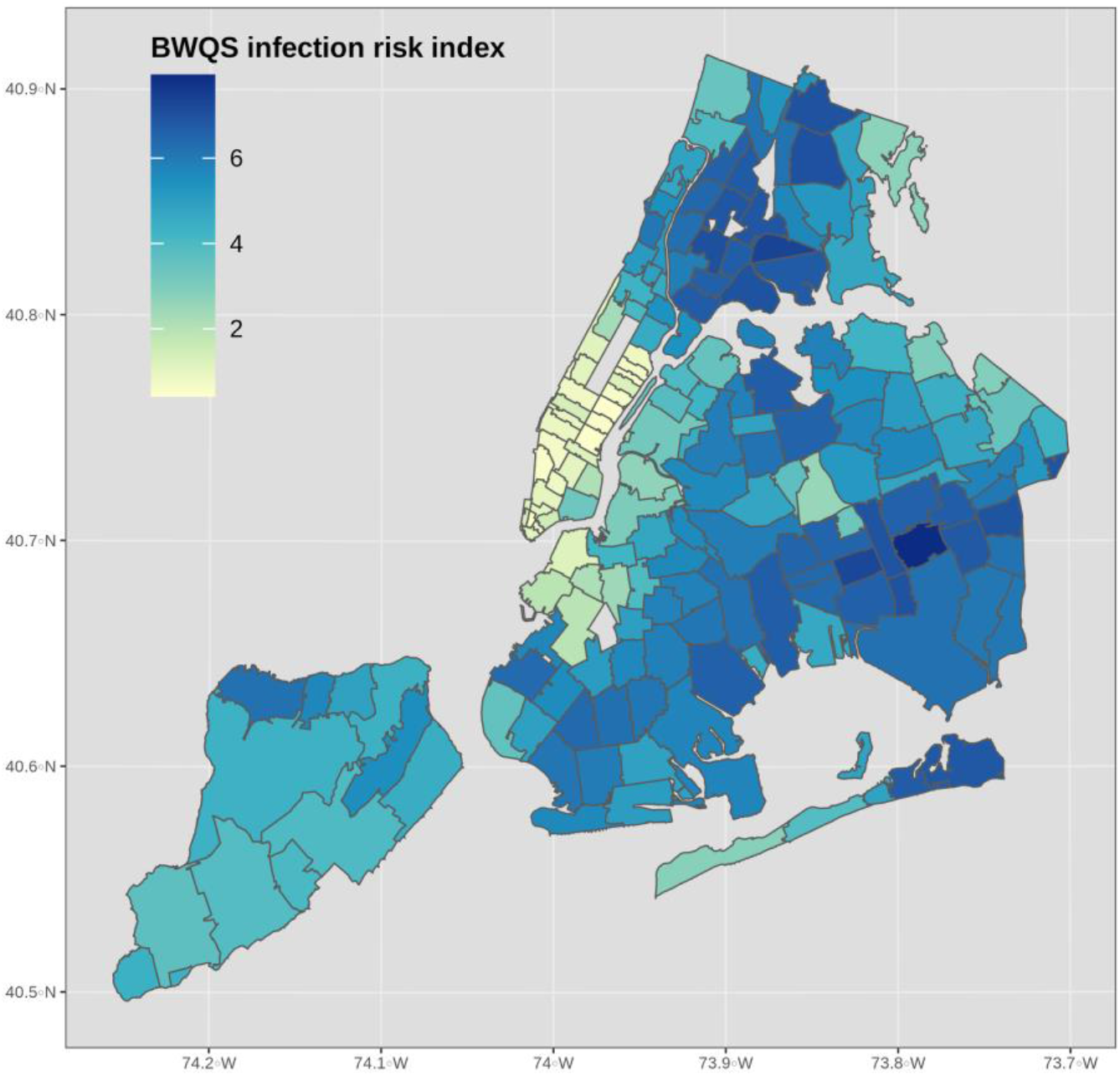
Map of NYC COVID-19 BWQS infection risk index. Unit of analysis is ZCTA (n=177).

**Fig. 4.**
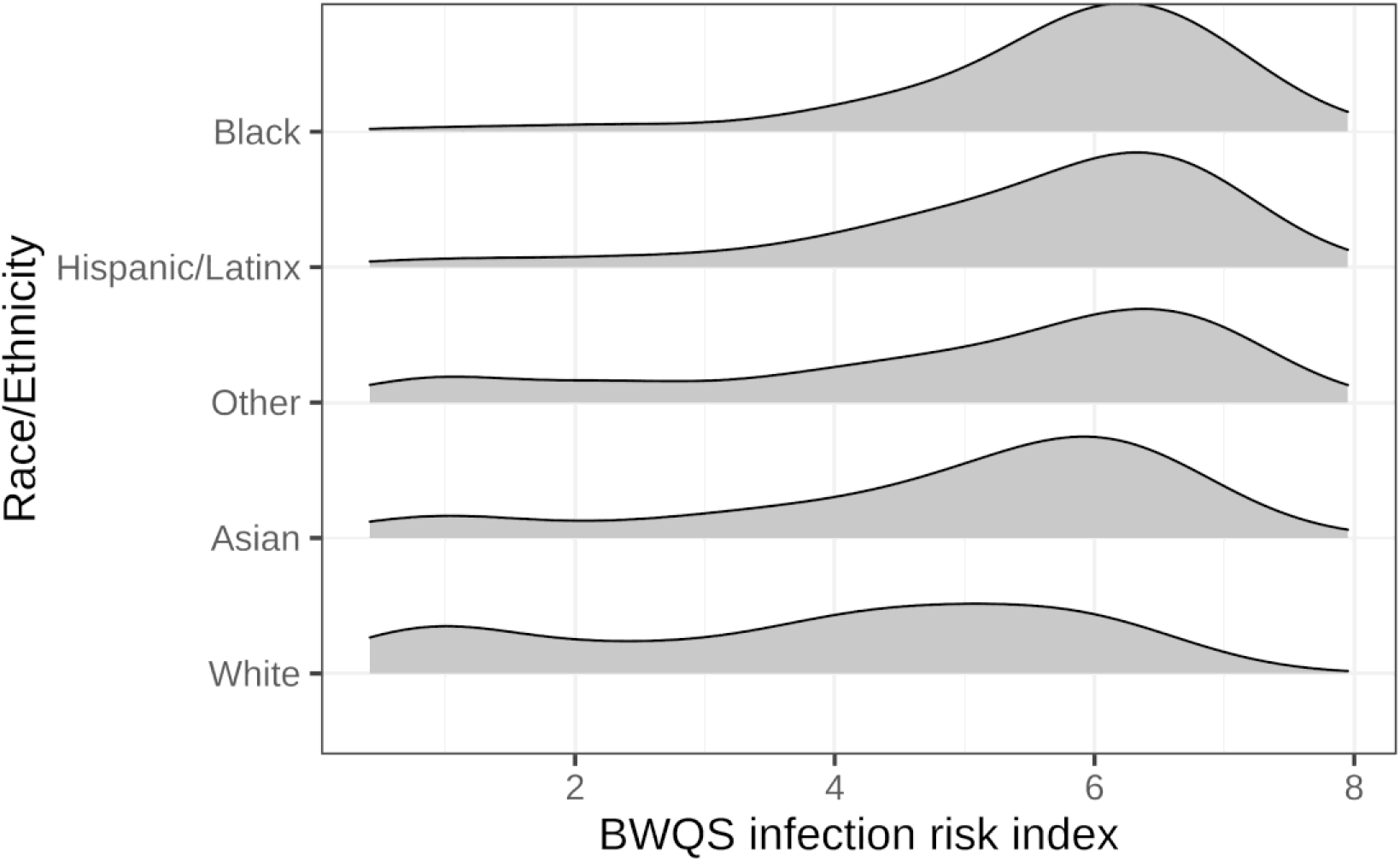
Distribution of BWQS infection risk index by race/ethnicity of ZCTA residents. Infection risk varied by race/ethnic categories according to the 2018 ACS. Density plots are population-weighted to demonstrate the relative abundance of representation according to ZCTAs and their corresponding BWQS infection risk index and ordered by the population-weighted mean index.

### Capacity to social distance

We found that capacity to social distance appears lower in higher neighborhood infection risk areas, as indicated by the most important variables in our neighborhood infection risk analysis. To assess whether or not this was true using longitudinal data, we decided to model differences in subway utilization by UHFs in NYC. We only included UHFs with the most consistent data quality and that had subways present (**Supplemental Figure 2**). In order to identify the proper functional form of our nonlinear model, we fit it on the mean sigmoidal decay of subway utilization across all of NYC (**Supplemental Figure 3**). We then compared this model to an interaction model for UHF-level population-weighted BWQS index (**Figure 5**). A partial F-test demonstrated that a model with an interaction term for BWQS index categories (above versus below the median) was a significantly better fit than one without the interaction term (p<0.0001).

**Fig. 5.**
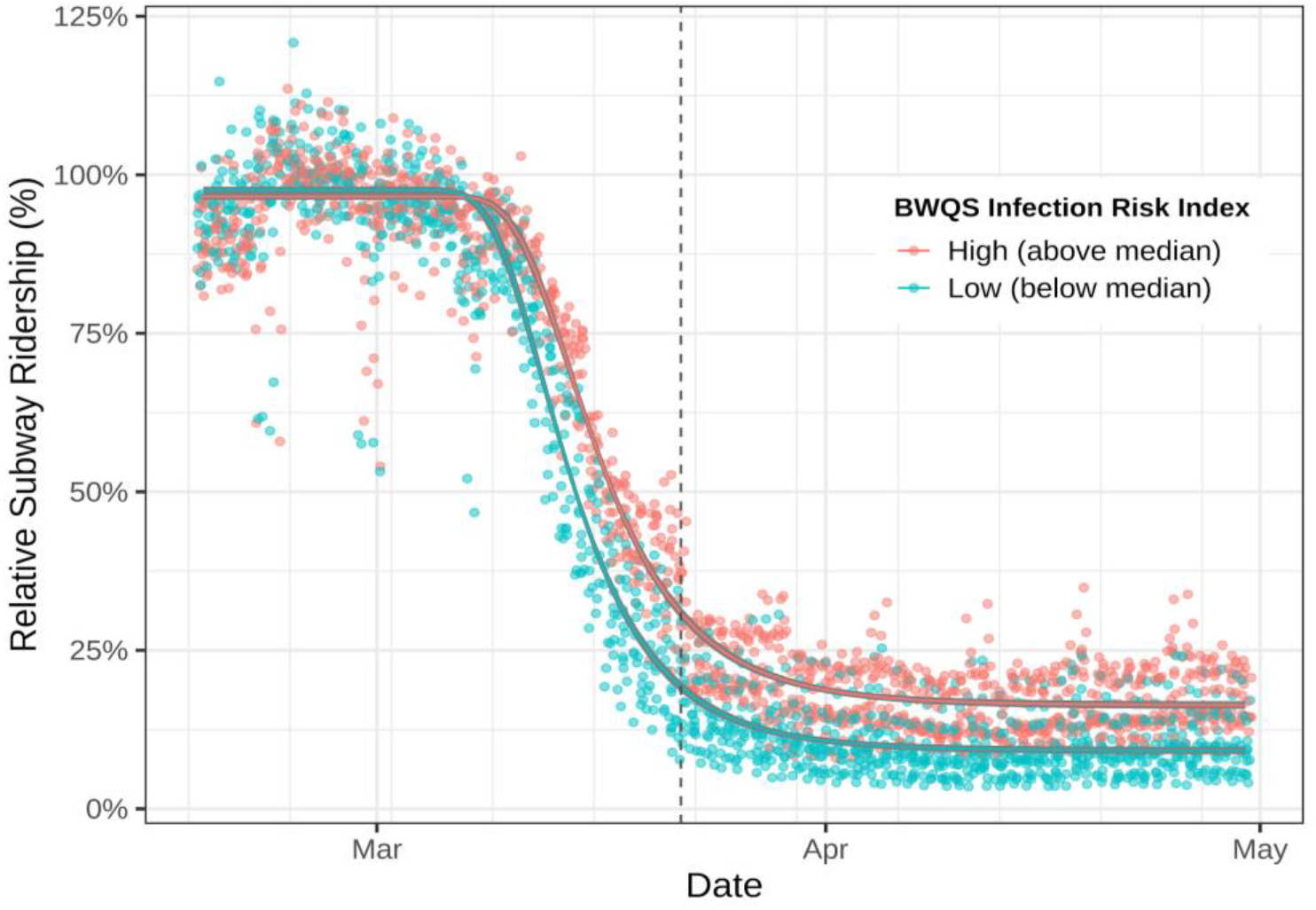
Subway ridership trends by population-weighted BWQS infection risk index at the United Hospital Fund neighborhood level. The nonlinear model was fitted using a generalized Weibull equation with two curves: high (above median) and low (below median) BWQS infection risk index at the UHF neighborhood level (n=36). Daily subway ridership is relative to 2015-2019. Dashed line represents the start of NYS on PAUSE social distancing policies. Ridership is shown between February 16, 2020 to April 30, 2020.

The interaction model indicates that there is no difference between slopes for the high (−5.6% per day; 95% CI: −5.9, −5.3%) versus low (−6.3% per day; 95% CI: −6.7, −5.9%) infection risk areas (**Table 1**). However, the lower asymptote of subway utilization under social distancing policies is higher for high infection risk (16%; 95% CI: 15.3, 16.7%) areas compared to low risk infection risk areas (9.6%; 95% CI: 8.8, 10.1%). This implies that high risk and low risk areas had similar relative rates of decreased subway utilization upon news of the pandemic, i.e. school closures, etc. However, high risk neighborhoods had a higher relative use of the subway system after official social distancing policies (NYS on PAUSE) went into effect.

**Table 1.** Coefficients from nonlinear regression of BWQS infection risk and subway ridership.

|  | High Infection Risk |  | Low Infection Risk |  |  |
| --- | --- | --- | --- | --- | --- |
|  | Coefficient | 95% CI | Coefficient | 95% CI | P value |
| **Lower Asymptote** | 16% | 15.3, 16.7% | 9.6% | 8.8, 10.1% | <0.001 |
| **Slope** | -5.6% per day | -5.9, -5.3% | -6.3% per day | -6.7, -5.9% | 0.52 |
| Lower Asymptote and Slope are parameters from a nonlinear model with generalized Weibull equation and two curves for high (above median) and low (below median) BWQS infection risk index at the UHF neighborhood level. |  |  |  |  |  |

#### Mortality related to neighborhood infection risk index

There were 16,289 COVID-related deaths across 177 ZCTAs by May 23, 2020. Results from the negative binomial model show an association between the ZCTA BWQS infection risk index and cumulative COVID mortality incidence (**Table 2**). This regression model employed a spatial filtering approach to account for potential spatial autocorrelation at the ZCTA level. We found that each unit increase in the BWQS infection risk index is associated with a 21% increased risk of COVID-related mortality (Relative Risk: 1.21; 95% CI: 1.16, 1.26) when adjusting for the proportion of the population aged 65+ and accounting for spatial dependence. There was non-significant spatial autocorrelation in the residuals (Moran’s I: 0.07, p value: 0.064).

**Table 2.** Results from a negative binomial regression of BWQS infection risk index and COVID-19 cumulative mortality proportion at the ZCTA level.

| Term | Relative Risk (95% CI) | P value |
| --- | --- | --- |
| BWQS infection risk index | 1.21 (1.16, 1.26) | <0.001 |
| Proportion 65 plus (z-score) | 1.03 (0.95, 1.11) | 0.153 |
| Eigenvector of spatial element | 0.35 (0.16, 0.76) | <0.001 |

## Discussion

We conducted a study using publicly-available data to identify the role of neighborhood social disadvantage on cumulative COVID-19 infections and COVID-19-related mortality. The neighborhood infection risk index was also used to understand differences in social distancing, as measured by subway ridership. In creating our neighborhood infection risk index, we found that a combination of social variables, indicative of social disadvantage, is associated with cumulative infections and mortality. Black and Hispanic/Latinx communities are overrepresented in high infection risk neighborhoods, and Whites are overrepresented in low infection risk neighborhoods, which may represent structural forms of racism. When examining differences in capacity to socially isolate, we found that high risk neighborhoods had higher subway ridership during NYS-mandated social distancing. Finally, our neighborhood infection risk index is also associated with cumulative COVID-19 mortality at the ZCTA level. This implies that the same social factors that inform increased disease risk are also associated with severe outcomes, either directly or through intermediates.

A growing body of literature is examining the greater impact of COVID-19 on communities of color. As some have noted, COVID-19 is not creating new health disparities, but exacerbating those that already exist^3^. A recent investigation found that county and ZCTA area-based socioeconomic measures, specifically using crowding, percent POC, and a measure of racialized economic segregation, were useful in identifying higher COVID-19 infections and mortality in Illinois and New York^18^. Work on COVID-19 mortality in Massachusetts has found excess death rates for areas of higher poverty, crowding, proportion POC, and racialized economic segregation^19^. Similarly, researchers have begun to identify counties that are particularly susceptible to severe COVID-19 outcomes using a combination of biological, demographic, and socioeconomic variables^20^. They identify areas with high population density, low rates of health insurance, and high poverty as particularly at risk. However, a stated limitation of this work is that many of these variables are interrelated.

Our study has many strengths. First, we acknowledge and address the strong interrelation of social variables by using a data-driven method for modeling mixtures of exposures: BWQS. By using this method, we create a composite index that captures the combined effect of the constituent variables. This process is also supervised, meaning that the variables are not weighted equally in the composite index, but instead the approach empirically learns their individual contributions to explaining the outcome. Others have addressed the multicollinearity of social determinants with the use of dimensionality reduction techniques such as principal components analysis (PCA) in the case of the neighborhood deprivation index^21^. However, traditional PCA only considers correlations between SES variables, whereas a supervised method captures features most relevant for the outcome. Second, our approach largely relies on ACS data, which is available across the USA, and may allow for the identification of other communities nationwide that are particularly vulnerable to future outbreaks or even other novel respiratory pathogens. Third, we explicitly excluded race and ethnicity from the creation of the index because we were more interested in identifying social processes that may facilitate infection risk, rather than those that may imply biological or behavioral explanations to health disparities^22^. The theory underlying these relationships is that structural racism sorts POC into areas of high disadvantage, and those structural forms of disadvantage facilitate pathogen spread. To demonstrate this, we employed the index to understand neighborhood differences in capacity to social distance. This finding provides additional evidence that low-income communities and communities of color may be less able to socially distance^6^. Fourth, our spatial analysis of COVID-19-mortality shows that the BWQS risk index may not only be useful in identifying infection risk, but also risk of severe outcomes. Finally, our data sources and analysis code are publicly-available, meaning that others can 1) reproduce these analyses, 2) expand on the work by assessing different modeling strategies and 3) assess the utility in other parts of the country.

This study also has notable limitations. First, we were unable to identify a measure of multigenerational housing at the ZCTA level, which may represent a pathway for infection, and potentially severe disease. Second, by not including race in our models, we may be missing an opportunity to tune these models to the impacts of interpersonal and structural forms of racism^23^. Third, early testing data in NYC was largely limited to hospitalized individuals, therefore those with more severe disease^9^. Consequently, ZCTA infection data may be confounded by the distribution of factors that drive disease severity. We addressed this by adjusting our BWQS regression for the amount of overall testing per ZCTA. Relatedly, for our spatial analysis of COVID-mortality, we were unable to access a ZCTA-level measure of chronic diseases. Since communities of color have higher rates of chronic disease at younger ages^24^, and chronic diseases increase the likelihood of severe COVID-19 outcomes, this is an important challenge. However, because social disparities are a major contributor to differences in the chronic conditions that increase the likelihood of severe disease, we did not want to adjust for a causal intermediate. Instead, we adjust for spatial autocorrelation to account for residual risk factors that are more similar in nearby neighborhoods. Fourth, we use pre-pandemic social variables derived from the 2018 ACS and thus do not directly account for variation in mobility^25^. However, this should be captured, in part, by median income and other measures of affluence in our BWQS index. Fifth, our analysis of public transit only utilized data from subway turnstiles, but not bus ridership. Although buses are an important form of transit in NYC, especially in the outer parts of the boroughs, the MTA does not provide time-varying ridership data. Further, buses were made free during the pandemic, so accurate ridership data are likely unavailable to the NYC government as well^26^. Finally, an unfortunate potential consequence of creating a neighborhood risk index is the possibility of stigmatization of neighborhoods with high risk index values^22^. This is not our intention, and hopefully not the effect, as our goal is to identify social factors that facilitate viral spread, and demonstrate that current public health guidance is not equally observable by all populations. Therefore, it is up to policymakers and practitioners to identify those populations and design/implement interventions accordingly.

## Conclusion

In this study, we created a neighborhood measure of social disadvantage that is specifically tuned to the impacts of COVID-19 infections and mortality and we show that this measure is associated with the capacity to socially distance, which may represent an important pathway for COVID-related health disparities. This is an important area of investigation given the large toll that COVID-19 has had, and will likely continue to have unless action is taken, on disadvantaged communities of color in NYC and elsewhere.

## Methods

### Data sources and cleaning

#### SARS-CoV-2 testing and COVID-19 mortality data

The New York City Department of Health and Mental Hygiene (NYC DOHMH) has been publicly releasing daily testing data (positive and total tests) at the patient’s home ZIP Code Tabulation Area (ZCTA) level since April 1, 2020, and COVID-19 related mortality data since mid-May, both available on GitHub^27^. The NYC DOHMH utilizes modified ZCTA geographies, designed to still be mergeable to the Census Bureau ZCTA designations. Our analyses relied on pre-pandemic demographic data to describe variation in neighborhood-level disease burden after much of the community had potential for exposure. Since spatiotemporal patterns in infection risk were highly variable at the beginning of the pandemic in relation to many independent viral introductions within NYC^28^, we estimated cumulative infections on May 7, 2020, four weeks after NYC’s peak infection period. We estimated time from symptom onset to death as 16 days^29^. Therefore we chose May 23, 2020 for our cumulative COVID-19 mortality analysis. This analysis is not human subjects research as it did not include any intervention or interaction with individuals or any identifiable private information.

#### Census data

We downloaded the Census Bureau’s 2018 American Community Survey (ACS) data via the *tidycensus* R package^30^. Data were collected for the 177 ZCTAs in NYC. Variables included: the total population, number of households, median income, median rent, health insurance status, unemployment, individuals at or below 150% of the federal poverty level, race and ethnicity, industry of employment, and mode of transportation to work. A full list of variables are provided in ***Supplemental Table 1***. We created a proxy for proportion in essential worker positions using industry of employment variables. This estimate of essential workers was a sum of those who reported employment in the agricultural, construction, wholesale trade, transportation and utilities, and education/healthcare industries, divided by the total working-age population. To account for teachers mostly working from home, and healthcare workers being essential, we included only half of the education/healthcare industry respondents. From these data we also estimated the average household size by dividing the total population by the number of households. We utilize race and ethnicity according to the following categories: Non-Hispanic Asian, Non-Hispanic Black, Non-Hispanic White, Hispanic/Latino of any race, and aggregate all other races into Other.

#### Residential buildings and food access data

We calculated the volume of residential space by merging datasets the NYC building footprints dataset^31^ and merged it with the Primary Land Use Tax Lot Output (PLUTO) dataset^32^. We divided residential volume by total population to calculate mean residents per residential volume, a metric of residential population density. Food access was used as a measure for the likelihood that individuals need to leave their neighborhoods for basic necessities. We estimated food access using data from New York State’s Open Data portal for Retail Food Stores^33^. Businesses were restricted to J, A, and C establishment code designations in order to identify those most likely to provide fresh foods and produce, and then manually removed any business names that indicated being a corner store or pharmacy, or primarily selling alcohol/tobacco. We spatially joined the point locations to our ZCTA shapefile and divided by the total Census population to calculate a ‘grocers per 1,000 people’ variable as a proxy for food access.

#### Mobility and transit data

The Metropolitan Transit Authority (MTA) of NYC releases subway utilization data on a weekly basis^34^. These data include the number of entrances and exits per station. For each day and geographic area, we summed all system entrances and exits.To account for typical usage of the subway on each month and day of the week, we divided the total turnstile count for each day and area by the median daily count on the same day of the week within the same month throughout the period 2015-2019.

### Quantitative Analyses

#### Cross-sectional Neighborhood Infection Risk Index

Socioeconomic variables are known to be closely correlated with one another, which is a challenge to model fitting and interpretation of the underlying latent relationship. To address these challenges, we develop a weighted combination of socioeconomic variables to explain the cumulative number of COVID-19 cases per ZCTA using Bayesian weighted quantile sums regression (BWQS)^35^. BWQS distinguishes two groups of predictors. In one group, which comprises our socioeconomic variables, the predictors are transformed into decile ranks to limit the influence of outliers, and the coefficients are forced to lie in [0, 1] and sum to 1 with a uniform Dirichlet prior. We included a large candidate list of socioeconomic variables in the BWQS that could represent some of the underlying infection dynamics attributable to socioeconomic disadvantage. They included selected demographic variables collected from the 2018 ACS, as well as derived variables such as population density (persons per square foot of the ZCTA) and residential population density (persons per cubic foot of ZCTA residential volume). Our final list of variables was based on an iterative process according to: 1) maximizing model fit, measured by the widely applicable information criterion (WAIC), 2) removing one variable when bivariate correlations were high (|*τ*| ≥ 0.9), and 3) our understanding of underlying social processes in relation to infectious disease. The other group of variables in a BWQS regression are the covariates, which in our case consist solely of the population-adjusted total number of tests administered per ZCTA. We included this to account for variation in disease surveillance. The predictor is untransformed and the coefficient is less constrained, using a normal prior with mean 0 and SD 100. A negative-binomial distribution is used for the dependent variable: the cumulative number of positive SARS-CoV-2 tests per 100,000 people. The resulting weighted index was our neighborhood infection risk index.

We visualized the distribution of the neighborhood risk scores by self-reported race/ethnicity as per the ACS categories and total population. We also separate the neighborhood risk scores into three categories: below the 25th percentile, between the 25th and 75th percentiles, and above the 75th percentile. Populations were aggregated by race/ethnicity and then divided by the total population of the associated ZCTAs.

#### Capacity to Social Distance

Our BWQS model uses cross-sectional data to create an infection risk index, but we wanted to assess the degree to which those differences in infections were explained longitudinally by inability to socially isolate/distance. We utilized MTA transit data as a proxy for social distancing since public transit may reflect conditions that contribute to greater exposure risks. Subway stations are in a fraction of NYC ZCTAs, and individuals often traverse ZCTAs to get to a station, so we aggregated subway utilization to 42 United Hospital Fund (UHF) neighborhoods. UHF neighborhoods are composed of adjacent ZCTAs approximating community districts. Aberrantly low utilization observations (<10%) in February and early March 2020 were removed when explained by planned weekend service changes - specifically those in low subway density areas. We computed a population-weighted BWQS index per UHF.

We modeled change in relative subway usage leading up to, and during, the NYS on PAUSE period. Relative subway utilization is a proportion, therefore the transition from business-as-usual to social distancing roughly followed a sigmoidal decay. A mean nonlinear response can be modeled by nonlinear least squares when a functional form is specified, as implemented by the *drc* R package^36^. We utilized a generalized Weibull formula, which took the following functional form:

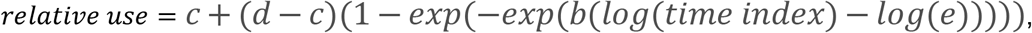

where *c* is the lower asymptote, *d* is the upper asymptote, *b* is the slope, *time index* is transformation of the date as an integer, *e* is the inflection point of the function, and *relative use* is the proportion of subway ridership. The model accommodates curve fitting with interaction terms to identify differences in model fit per group. For ease of interpretation and visualization, this was utilized to assess differences for high (above the median) versus low (below the median) BWQS index neighborhoods. We sought to identify any differences in slope (b) and the lower asymptote (c) as indicative of differences in the ability to socially isolate. An F-test was used to compare a naive model (without considering the BWQS index) to a model with interaction by high versus low BWQS index.

#### Neighborhood infection risk and mortality

Given high COVID-related mortality in disadvantaged communities, we wanted to assess if our measure of neighborhood infection risk was also associated with cumulative COVID mortality by total population. To do so, we employed a negative binomial model, regressing ZCTA-level COVID mortality on the BWQS infection risk index, adjusting for the proportion of the population that was greater than or equal to 65 years old. In order to adjust for spatial autocorrelation, and thus unmeasured spatial confounding, we employed a spatial filtering approach whereby we identify the eigenvector associated with spatial autocorrelation (as measured by Moran’s I), and explicitly adjusted for those values in the negative binomial regression^37,38^. The goal, then, was to “filter out” spatial autocorrelation from the residuals. Negative binomial models were implemented with the *MASS* package, supplemented with spatial functions from the *spdep* and *spatialreg* packages^39,40^.

### Mapping and coding

Geoprocessing and visualization of spatial data were conducted with the *sf* package in R^41^. All analyses were conducted in R version 3.6.2^42^ and the code is available via GitHub.

## Data Availability

All data were derived from public use datasets via government websites. All analytic code, including download procedures, are available to the public in GitHub.

https://github.com/justlab/COVID_19_admin_disparities/tree/v1

## Data Availability

All data were derived from public use datasets via government websites. All analytic code, including download procedures, are available to the public in GitHub at https://github.com/justlab/COVID_19_admin_disparities/tree/v1

## Acknowledgements

This work was supported by grant UL1TR001433 and P30ES023515. DC is funded by NIH T32HD049311. Thanks to Sebastian Rowland for his thoughtful comments on a draft.

## Contributions

DC and ACJ conceptualized the study and DC drafted the manuscript. DC conducted all analyses with statistical support from EC and NFP. ACJ, EC, and ND provided feedback on design and analysis. The Bayesian Weighted Quantile Sum regression was designed and implemented by EC and NFP, with the log link function implemented by NFP. KA developed the procedures and indices for relative subway utilization. JR ingested DOHMH data. All authors reviewed and approved the manuscript.

## Supplemental materials

**Supplemental Table 1:**
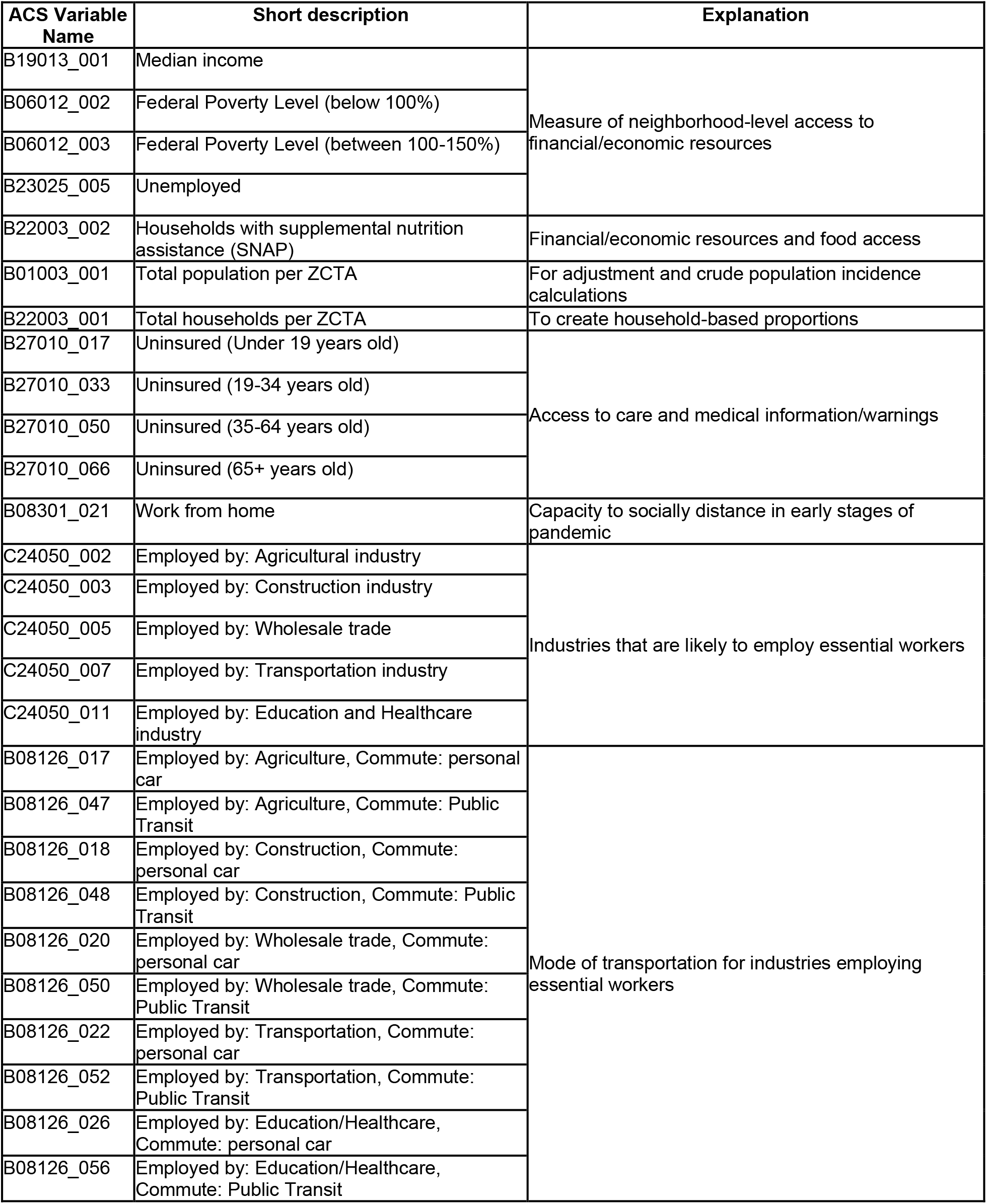
2018 American Community Survey (ACS) 5-year estimate variables collected via *tidycensus* R package for ZIP code tabulation area (ZCTA) units.

**Supplemental Fig. 1.**
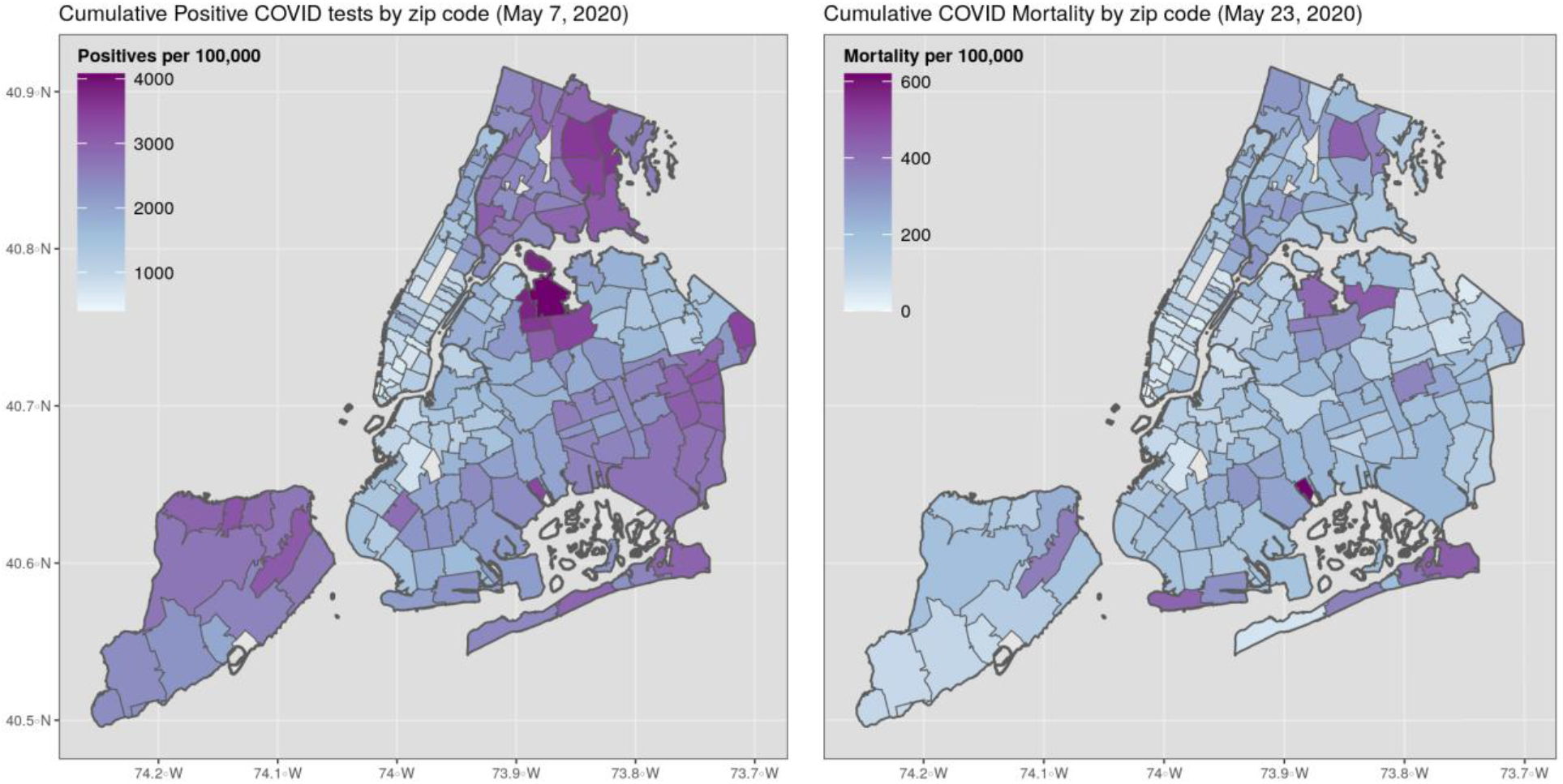
Cumulative infections and COVID mortality per 100,000 by ZCTA as of May 7, 2020 and May 23, 2020 respectively.

**Supplemental Fig. 2.**
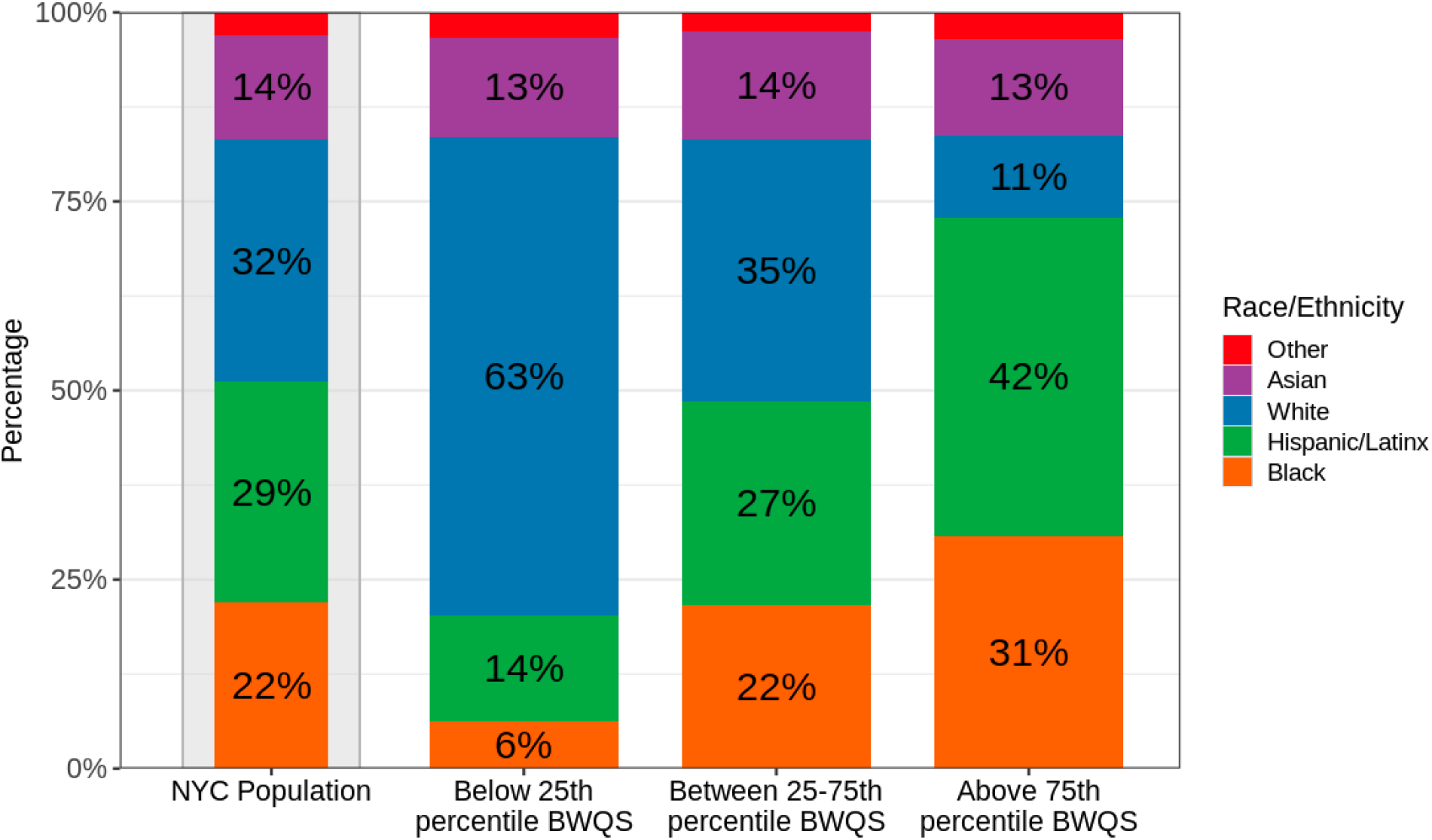
Race/ethnic composition of ZCTAs that fall within various quantiles of the BWQS infection risk index. Race/ethnic data from the 2018 ACS. NYC total demographic breakdown provided as reference.

**Supplemental Table 2:** Kendall’s tau correlations for social determinants with per capita cumulative SARS-CoV-2 infection incidence as of May 7, 2020 by NYC ZCTA.

| Variable | Correlation | P value |
| --- | --- | --- |
| 1/median income | 0.317 | <0.0001 |
| Not insured (%) | 0.231 | <0.0001 |
| Unemployed (%) | 0.296 | <0.0001 |
| 1/grocers per 1000 people | 0.261 | <0.0001 |
| Essential workers (%) | 0.544 | <0.0001 |
| Essential workers commuting via public transit (%) | 0.174 | 0.0001 |
| Essential workers commuting via car (%) | 0.493 | <0.0001 |
| People who do not work from home (%) | 0.454 | <0.0001 |
| Population density according to housing volume (people per cubic foot) | 0.096 | 0.058 |
| Average household size (#) | 0.467 | <0.0001 |

**Supplemental Fig. 3.**
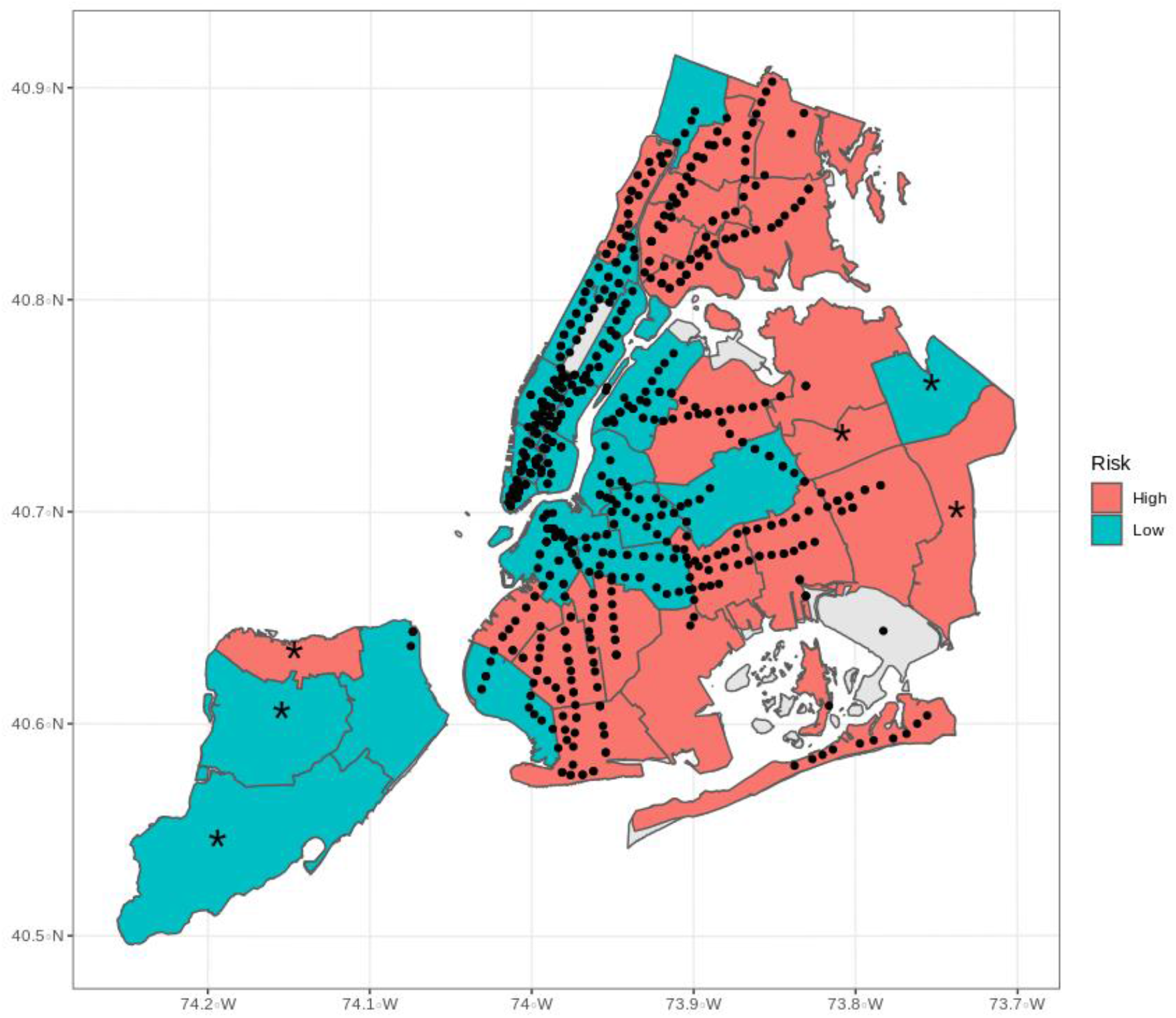
UHF neighborhoods by population-weighted neighborhood infection risk index. Six UHF neighborhoods excluded from the subway ridership analysis are indicated by asterisks. Dots represent subway stations with available data. UHF neighborhoods are colored as high (above median) or low (below median) BWQS infection risk index.

**Supplemental Fig. 4.**
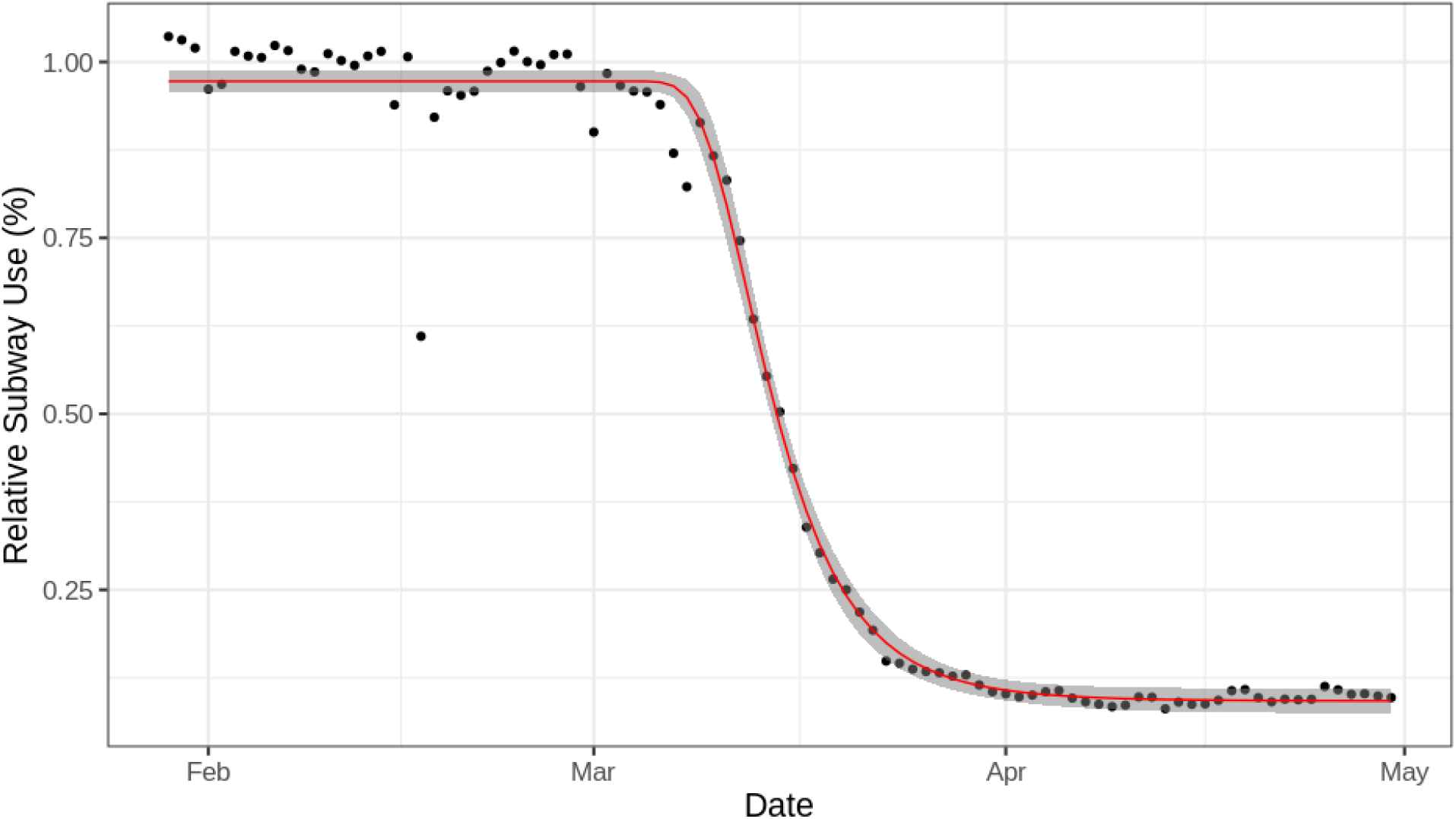
Fit of the generalized Weibull equation curve on the citywide mean of UHF-level ridership. Outlier on February 15th, a national holiday. Values above 1 indicate above-average ridership per our comparison with 2015-2019.

## References

1. Onder, G., Rezza, G. & Brusaferro, S. Case-Fatality Rate and Characteristics of Patients Dying in Relation to COVID-19 in Italy. JAMA 323, 1775–1776 (2020).

2. New York State Department of Health. New York State on PAUSE. Department of Health https://coronavirus.health.ny.gov/new-york-state-pause (2020).

3. Dorn, A. van, Cooney, R. E. & Sabin, M. L. COVID-19 exacerbating inequalities in the US. The Lancet 395, 1243–1244 (2020).

4. Gold, J. A. W. Characteristics and Clinical Outcomes of Adult Patients Hospitalized with COVID-19 — Georgia, March 2020. MMWR Morb. Mortal. Wkly. Rep. 69, (2020).

5. Webb Hooper, M., Nápoles, A. M. & Pérez-Stable, E. J. COVID-19 and Racial/Ethnic Disparities. JAMA (2020) doi:10.1001/jama.2020.8598.

6. Yancy, C. W. COVID-19 and African Americans. JAMA (2020) doi:10.1001/jama.2020.6548.

7. Centers for Disease Control & Prevention. CDC Health Disparities and Inequalities Report — United States, 2013. https://www.cdc.gov/mmwr/pdf/other/su6203.pdf (2013).

8. Garg, S. Hospitalization Rates and Characteristics of Patients Hospitalized with Laboratory-Confirmed Coronavirus Disease 2019 — COVID-NET, 14 States, March 1-30, 2020. MMWR Morb. Mortal. Wkly. Rep. 69, (2020).

9. NYC DOHMH. Age-adjusted rates of lab-confirmed COVID-19. Age-adjusted rates of lab-confirmed COVID-19 https://www.1.nyc.gov/assets/doh/downloads/pdf/imm/covid-19-deaths-race-ethnicity-05142020-1.pdf (2020).

10. Krieger, N. Discrimination and health inequities. Int. J. Health Serv. 44, 643–710 (2014).

11. Bailey, Z. D. et al. Structural racism and health inequities in the USA: evidence and interventions. The Lancet 389, 1453–1463 (2017).

12. Acevedo-Garcia, D. Residential segregation and the epidemiology of infectious diseases. Soc. Sci. Med. 51, 1143–1161 (2000).

13. Acs, G. & Loprest, P. J. Job Differences by Race and Ethnicity in the Low-Skill Job Market. 6 https://www.urban.org/sites/default/files/publication/30146/411841-Job-Differences-by-Race-and-Ethnicity-in-the-Low-Skill-Job-Market.PDF (2009).

14. Berchick, E. R., Barnett, J. C. & Upton, R. D. Health Insurance Coverage in the United States: 2018. 44 https://www.census.gov/content/dam/Census/library/publications/2019/demo/p60-267.pdf (2019).

15. Murray, C. J. L. et al. Eight Americas: Investigating Mortality Disparities across Races, Counties, and Race-Counties in the United States. PLoS Med. 3, e260 (2006).

16. Guzman, S. Multigenerational Housing on the Rise, Fueled by Economic and Social Changes. https://www.aarp.org/ppi/info-2019/multigenerational-housing.html (2019) doi:10.26419/ppi.00071.001.

17. Walker, R. E., Keane, C. R. & Burke, J. G. Disparities and access to healthy food in the United States: A review of food deserts literature. Health Place 16, 876–884 (2010).

18. Chen, C., Jarvis T. & Krieger, N. Revealing the unequal burden of COVID-19 by income, race/ethnicity, and household crowding: US county vs. ZIP code analyses. (2020).

19. Chen, J., Waterman, P. & Krieger, N. COVID-19 and the unequal surge in mortality rates in Massachusetts, by city/town and ZIP Code measures of poverty, household crowding, race/ethnicity,and racialized economic segregation. Harvard Center for Population and Development Studies Working Paper Series https://www.hsph.harvard.edu/population-development/research/working-papers/harvard-pop-center-working-paper-series/ (2020).

20. Chin, T. et al. U.S. county-level characteristics to inform equitable COVID-19 response. http://medrxiv.org/lookup/doi/10.1101/2020.04.08.20058248 (2020) doi:10.1101/2020.04.08.20058248.

21. Messer, L. C. et al. The Development of a Standardized Neighborhood Deprivation Index. J. Urban Health 83, 1041–1062 (2006).

22. Chowkwanyun, M. & Reed, A. L. Racial Health Disparities and Covid-19 — Caution and Context. N. Engl. J. Med. 0, null (2020).

23. Geronimus, A. T., Hicken, M., Keene, D. & Bound, J. “Weathering” and age patterns of allostatic load scores among blacks and whites in the United States. Am. J. Public Health 96, 826–833 (2006).

24. Gee, G. C., Hing, A., Mohammed, S., Tabor, D. C. & Williams, D. R. Racism and the Life Course: Taking Time Seriously. Am. J. Public Health 109, S43-S47 (2019).

25. Quealy, K. The Richest Neighborhoods Emptied Out Most as Coronavirus Hit New York City. The New York Times (2020).

26. Guse, C. NYC local buses a free ride for all during coronavirus outbreak. nydailynews.com https://www.nydailynews.com/coronavirus/ny-coronavirus-mta-nyc-buses-free-outbreak-20200320-a24jq7ksrzcpvgq7e2dyzq2uqy-story.html (2020).

27. NYC DOHMH. NYC DOHMH Coronavirus Data. GitHub https://github.com/nychealth/coronavirus-data (2020).

28. Gonzalez-Reiche, A. S. et al. Introductions and early spread of SARS-CoV-2 in the New York City area. Science (2020) doi:10.1126/science.abc1917.

29. CDC. Coronavirus Disease 2019 (COVID-19). Centers for Disease Control and Prevention https://www.cdc.gov/coronavirus/2019-ncov/hcp/planning-scenarios.html (2020).

30. Walker, K. tidycensus: Load US Census Boundary and Attribute Data as ‘tidyverse’ and ‘sf’-Ready Data Frames. (2020).

31. NYC Open Data. NYC Building Footprints. NYC Open Data https://data.cityofnewyork.us/Housing-Development/Building-Footprints/nqwf-w8eh (2020).

32. NYC Department of Planning. PLUTO and MapPLUTO. https://www.1.nyc.gov/site/planning/data-maps/open-data/dwn-pluto-mappluto.page (2020).

33. NYS Food Safety. Retail Food Stores. New York State Open Data https://data.ny.gov/Economic-Development/Retail-Food-Stores/9a8c-vfzj (2019).

34. NYC MTA. Metropolitan Transit Authority Information. Developer Data Downloads http://web.mta.info/developers/download.html (2020).

35. Colicino, E., Pedretti, N. F., Busgang, S. A. & Gennings, C. Per- and poly-fluoroalkyl substances and bone mineral density: Results from the Bayesian weighted quantile sum regression. Environ. Epidemiol. 4, e092 (2020).

36. Ritz, C., Baty, F., Streibig, J. C. & Gerhard, D. Dose-Response Analysis Using R. PLOS ONE 10, e0146021 (2015).

37. Griffith, D. A. & Peres-Neto, P. R. Spatial modeling in ecology: the flexibility of eigenfunction spatial analyses. Ecology 87, 2603–2613 (2006).

38. Griffith, D. A. Some robustness assessments of Moran eigenvector spatial filtering. Spat. Stat. 22, 155–179 (2017).

39. Bivand, R. S., Pebesma, E. & Gomez-Rubio, V. Applied spatial data analysis with R. (Springer, 2013).

40. Bivand, R. S. & Piras, G. Comparing Implementations of Estimation Methods for Spatial Econometrics. J. Stat. Softw. 63, 1–36 (2015).

41. Pebesma, E. Simple features for R: standardized support for spatial vector data. R J. 10, 439–446 (2018).

42. R Core Team. R: A Language and Environment for Statistical Computing. (R Foundation for Statistical Computing, 2019).

